# Assessing the validity of a self-reported clinical diagnosis of schizophrenia

**DOI:** 10.1101/2023.12.06.23299622

**Authors:** Grace E Woolway, Sophie E Legge, Amy Lynham, Sophie E Smart, Leon Hubbard, Ellie R Daniel, Antonio F Pardiñas, Valentina Escott-Price, Michael C O’Donovan, Michael J Owen, Ian R Jones, James TR Walters

## Abstract

**Background:** Diagnoses in psychiatric research can be derived from various sources. This study assesses the validity of a self-reported clinical diagnosis of schizophrenia.

**Methods:** The study included 3,029 clinically ascertained participants with schizophrenia or psychotic disorders diagnosed by self-report and/or research interview and 1,453 UK Biobank participants with self-report and/or medical record diagnosis of schizophrenia or schizoaffective disorder depressed-type (SA-D). We assessed positive predictive values (PPV) of self-reported clinical diagnoses against research interview and medical record diagnoses. We compared polygenic risk scores (PRS) and phenotypes across diagnostic groups, and compared the variance explained by schizophrenia PRS to samples in the Psychiatric Genomics Consortium (PGC).

**Results:** In the clinically ascertained sample, the PPV of self-reported schizophrenia to a research diagnosis of schizophrenia was 0.70, which increased to 0.81 when benchmarked against schizophrenia or SA-D. In UK Biobank, the PPV of self-reported schizophrenia to a medical record diagnosis was 0.74. Compared to self-report participants, those with a research diagnosis were younger and more likely to have a high school qualification (clinically ascertained sample) and those with a medical record diagnosis were less likely to be employed or have a high school qualification (UK Biobank). Schizophrenia PRS did not differ between participants that had a diagnosis from self-report, research diagnosis or medical record diagnosis. Polygenic liability r^2^, for all diagnosis definitions, fell within the distribution of PGC schizophrenia cohorts.

**Conclusions:** Self-report measures of schizophrenia are justified in research to maximise sample size and representativeness, although within sample validation of diagnoses is recommended.

## Introduction

Schizophrenia is characterised by positive, negative, and disorganised symptoms as well as cognitive deficits^1^ and has a lifetime prevalence of 0.32% worldwide^2^. In research studies, cases can be ascertained in various ways; for example, diagnoses can be based on self-report of a clinical diagnosis made by a health professional, electronic health records, or a combination of research interview and clinical note review. Methods combining diagnostic interviews and note reviews have been considered the gold-standard for defining cases in research^3^, but are resource intensive and often associated with ascertainment biases leading to unrepresentative samples^4, 5^. A recent study estimated that only one fifth of patients with schizophrenia are represented in randomised clinical trials in Finland and Sweden, most commonly due to patients being classed as ineligible as a result of somatic comorbidities, antidepressant/mood stabiliser use, substance use and suicide risk^6^. Relying on diagnoses obtained through diagnostic interviews also puts practical limitations on participation (e.g., only recruiting patients who are able to complete a lengthy interview), which may also affect the representation of the population of people with schizophrenia.

Diagnoses generated from medical records have been shown to be concordant with research interview diagnoses^7, 8^, with particularly high convergence seen in schizophrenia^9–11^. Ascertainment through records review overcomes some of the practical limitations for participation, but still hinders representation by relying on records typically from secondary care^3^ and thus under-represents patients who have not been admitted to hospital. One approach to improving the generalisability of research findings and increasing sample size could be to use self-reported diagnoses, circumventing the need for a labour-intensive research interview. However, the validity of self-reported diagnoses is likely to differ between psychiatric disorders, contexts, and cultures. Large-scale genomic datasets, such as 23andMe ^12–17^, UK Biobank ^13, 16, 18–21^ and the Million Veterans Programme ^16, 22^, have exploited self-report methods, but the reliability and validity of self-reported diagnoses is unclear^23^.

It is also unclear what impact different diagnostic methodologies have on the outcome of genetic studies^24^. In order to enhance power in Genome Wide Association Studies (GWAS) individuals with a self-reported diagnosis have been included ^13, 17^, and are likely to be increasingly so, despite some studies suggesting that individuals defined using minimal phenotyping approaches show genetic differences to participants who are strictly defined ^23, 25^. In one study, the effect sizes for schizophrenia polygenic risk scores (PRS) were reported to be smaller in samples where diagnoses are derived from electronic health records compared to clinically ascertained case-control research cohorts in the Psychiatric Genomics Consortium (PGC)^26^. However, analyses comparing samples from the Schizophrenia Working Group of the PGC found no differences in PRS across consensus DSM/ICD diagnosis (by psychiatrists), diagnostic interview, medical records, and mixed methods^27^. To our knowledge, there is no published research comparing a self-reported clinical diagnosis of schizophrenia from a health professional against a gold-standard research interview diagnosis.

In this study, we address this knowledge gap and assess whether a self-reported clinical diagnosis of schizophrenia is a valid approach to identify relevant individuals in genomic research.

## Methods

### Samples

Study participants came from a clinically ascertained sample which consisted of two Cardiff University cohorts, the National Centre for Mental Health (NCMH) and CardiffCOGS, and from the UK Biobank.

NCMH participants were recruited via health care services, voluntary organisations or via public advertisement^28^. Trained researchers administered a brief standardized assessment to gather demographic and clinical information. Participants reporting psychosis or mood symptoms were invited to take part in a research interview based on the Schedules for Clinical Assessment in Neuropsychiatry (SCAN)^29^. CardiffCOGS is a cohort recruited from the community, in-patient and voluntary sector mental health services across the UK^30^. All participants completed a SCAN-based research interview^29^ and underwent a case-note review. All participants from both studies were asked to provide a sample for DNA extraction and genetic analyses. NCMH and CardiffCOGS received approval from Health Research Authority and Wales Research Ethics Committee (REC) 2 (NCMH REC reference: 16/WA/0323), and Southeast Wales REC (CardiffCOGS REC reference: 07/WSE03/110). All participants provided written informed consent.

UK Biobank is a population-based UK cohort of around 500,000 participants, aged between 40-69 at recruitment^31^. Participants completed a range of assessments and provided a sample for genetic analysis. All participants provided written informed consent. Ethical approval was granted by the Northwest Multi-Centre Ethics Committee. This study was conducted under UK Biobank project number 13310.

### Diagnosis definitions

Table 1 provides an overview of the diagnosis definitions used in both the clinically ascertained sample and the UK Biobank alongside corresponding questions and variables from study assessments.

**Table 1.** Diagnosis variables used for self-report, research interview diagnosis and medical record diagnosis groups.

| Sample | Diagnosis type | Diagnosis subtype | Source | Diagnosis question/variable |
| --- | --- | --- | --- | --- |
| Clinically ascertained Sample | Self-report | Lifetime clinical diagnosis | Brief standardized assessment | “Has a doctor or health professional ever told you that you have any of the following diagnoses?” |
|  |  | Participant opinion | Brief standardized assessment | “What is your primary diagnosis, in your opinion?” |
|  |  | Current clinical diagnosis | Brief standardized assessment | “What is your primary diagnosis, in your clinician’s opinion?” |
|  | Research interview diagnosis |  | SCAN-based clinical interview | DSM-IV (1994), DSM-V (2013) and ICD-10 (1992) diagnoses |
| UK Biobank | Self-report |  | Mental health questionnaire | “Have you been diagnosed with one or more of the following mental health problems by a professional, even if you don't have it currently?” |
|  |  |  | Initial assessment | Verbal self-report of a mental health diagnosis during interview with a nurse |
|  | Medical record diagnosis |  | Primary care, hospital admission and death records | ICD-10 diagnosis |

*The diagnosis type and subtypes by diagnosis source and questions/variables used to derive definitions.*

### Self-reported diagnosis

In all samples, participants were asked whether a doctor or health professional had ever told the participant that they had a mental health diagnosis and given a list of psychiatric diagnoses to choose from (see Supplementary Figure 1 and 2). If the participant chose schizophrenia from the list or they verbally self-reported a schizophrenia diagnosis, they were assigned a schizophrenia self-report. See Table 1 for the subtypes of self-reported diagnoses in the clinically ascertained sample. A self-report of schizoaffective disorder was not analysed as it was not possible to differentiate between the subtypes.

### Research interview diagnosis

In the clinically ascertained sample, DSM-IV, DSM-V, and ICD-10 diagnoses were derived from a SCAN-based clinical interview and note review (where available). A diagnosis of schizophrenia was given in this study if either the DSM or ICD schizophrenia criteria was met. If participants met criteria for schizoaffective disorder depressed-type (SA-D), they were included alongside participants with schizophrenia. ‘Other psychotic disorders’ refer to the following diagnoses: psychosis not otherwise specified, schizophreniform disorder, delusional disorder, brief psychotic disorder, acute polymorphic disorder, and other psychotic illness.

### Medical record diagnosis

In UK Biobank, a medical record diagnosis of schizophrenia and SA-D were defined as a F20/F25.1 ICD-10 code from hospital admission records or death records, or an equivalent read code from primary care (Supplementary Table 1). Hospital records date back to 1997 for England, 1998 for Wales and 1981 for Scotland and contain coded data on admissions, operations, and procedures. Primary care data was obtained for approximately 45% of the UK Biobank cohort (∼230,000 participants).

Hospital admissions for schizophrenia were further subdivided into primary and secondary admissions. Primary ICD-10 codes represent conditions that caused the admission and secondary ICD-10 codes represent conditions that coexist at the time of admission, affect the treatment received, or develop after admission.

### Unaffected controls

Unaffected controls for the clinical samples were NCMH participants with no history of a mental health diagnosis and who were recruited through participants with a psychiatric diagnosis (e.g., a family member/partner) or via advertisements. Unaffected controls for the UK Biobank analyses consisted of participants in UK Biobank who did not have a psychotic disorder diagnosis.

### Phenotypic data

The phenotypes compared across diagnostic groups included sex, age at interview (in years), educational attainment, and employment status. Educational attainment was dichotomised to GCSE (General Certificate of Secondary Education) and above, usually achieved at 16 years upon completing high school, or below GCSE/no qualification consistent with previous research^32^ in addition to degree and no degree. Employment status was dichotomised to in current paid employment or not and restricted to participants under the age of 65 who did not report being retired.

### Genetic data

#### Clinically ascertained sample

The clinically ascertained participants were genotyped on the Illumina OmniExpress (Infinium OmniExpress-24 Kit), Illumina PsychArray (Infinium PsychArray-24 Kit) or Illumina GSA (Infinium Global Screening Array-24 Kit) genotyping platforms. Quality control and imputation using the Haplotype Reference consortium (HRC)^33^ was performed as part of the DRAGON-Data protocol^34^. Datasets containing participants from the clinically ascertained samples were restricted to those with the diagnoses described above and who did not carry a neurodevelopmental CNV^34^. These samples were combined with samples from 1000 Genomes European phase 3^35^ using PLINK v1.9 after restricting to overlapping SNPs. The 1000 Genomes sample was included to provide a population reference to allow studies using different arrays to be directly compared^36^. The following quality control exclusion criteria were subsequently applied to SNPs: minor allele frequency (MAF) < 0.05, genotyping rate < 0.05, and Hardy-Weinberg equilibrium p ≤ 10^−6^. Linkage disequilibrium-pruned SNPs (500 variant count window size, 20 variant count to shift the window at the end of each step, a pairwise r^2^ threshold of 0.2) were used to identify related individuals and to derive principal components (PC). One individual from each pair assumed to be duplicates (kinship coefficient > 0.98) or related (kinship coefficient > 0.1875) was removed at random. The first 5 PCs were used to perform multi-dimensional clustering to identify an ancestrally-homogenous subsample of individuals^37^ (n=1252). The first 5 PCs explained the majority of the variance in the principal components, adding additional PCs did not change the classifications. Individuals within a 90% threshold from the most central point were included for analyses. There were insufficient numbers of participants of non-European ancestries in NCMH and CardiffCOGS to allow us to analyse PRS in different ancestries.

#### UK Biobank

Imputed genetic data were provided by UK Biobank. Pre-imputation quality control and imputation have been described elsewhere^38^. Briefly, participants were assayed at the Affymetrix Research Services laboratory using the UK Biobank Axiom or UK BiLEVE Axiom purpose-built arrays. Imputation was completed using the HRC panel^33^. We applied additional quality control procedures using the same thresholds used in our clinically ascertained sample and detailed elsewhere^36, 39^. Genetic analyses were restricted to participants with a European ancestry, to mirror the clinically ascertained sample, using the method described above, see also Legge et al^39^.

### Polygenic risk scores

In the clinically ascertained sample and UK Biobank, PRSicev2^40^ was used to calculate PRS for schizophrenia using GWAS de-duplicated summary statistics that were derived separately from our clinical sample and UK Biobank^27^. Summary statistics underwent quality control^34^ and SNPs with MAF > 0.01 outside of the Major Histocompatibilty Complex region were used in the PRS analysis. PRS were calculated, using relatively independent SNPs (r^2^<0.1, within 500kb window), at a p-value threshold of 0.05^27^. Polygenic risk scores were standardised within samples prior to analysis.

### Analysis

In the clinically ascertained sample, positive predictive values (PPV) were used to assess the self-reported diagnosis from a health professional against the DSM/ICD research interview diagnosis in the same participants. We consider a research interview diagnosis of schizophrenia and schizoaffective disorder depressive-type (SA-D) together as there is evidence these two groups do not substantially differ with respect to genetic liability to schizophrenia^27, 41^. It was not possible to assess negative predictive values (NPV), sensitivity and specificity in the clinically ascertained sample due to the recruitment methods; participants were only approached to complete a SCAN-based research interview if they self-reported a mood or psychotic disorder diagnosis.

In the UK Biobank, PPV, NPV, sensitivity and specificity were used to assess how predictive a self-reported clinical diagnosis from a health professional was of a medical record diagnosis. We additionally scaled the PPV and NPV to the population point prevalence of schizophrenia (0.6%) (See Supplementary Note). We did not consider a medical record diagnosis of schizophrenia and SA-D together in the PPV analysis due to a very low prevalence of SA-D in the UK Biobank.

In both the clinically ascertained sample (NCMH participants only) and the UK Biobank, logistic regressions were used to test for phenotypic differences across the self-report-only and research interview diagnosis/medical record diagnosis groups. Year of birth and sex were included as covariates.

Similarly in both samples, logistic regressions were used to test for genetic differences in schizophrenia between the self-report-only and the research interview diagnosis/medical record diagnosis groups. In the UK Biobank sample, further logistic regressions were used to assess if schizophrenia PRS was associated with the number of times a diagnosis was reported, the number of admissions and type of admission (primary and secondary). These PRS analyses were covaried for first 5 PCs, array, age at assessment, and sex.

We compared the variance explained by PRS on the liability-scale (r^2^, assuming 1% lifetime risk) in schizophrenia case/control status in the clinically ascertained sample and UK Biobank, separated by diagnosis definitions, against the variances reported by other samples of European genetic ancestry in the PGC3 schizophrenia. The r^2^ values refer to the variance explained by the schizophrenia PRS in comparison to a covariates-only baseline model. In addition, we calculated the variance explained in schizophrenia case/control status by PRS for bipolar disorder^13^ and major depressive disorder^42^, which have been previously described^39^.

## Results

A total of 3,778 clinically ascertained participants (3,029 cases and 749 controls) and 502,541 UK Biobank participants (1,453 cases and 501,088 controls) were included in this study. In NCMH, a total of 1112 participants had schizophrenia or SA-D diagnosis, of which 654 had a self-report, 458 had a research diagnosis, and 365 had both. In the clinically ascertained sample (including NCMH and CardiffCOGS participants), 803 participants had a research diagnosis and 449 had exclusively a self-report diagnosis. The mean age of recruitment in the clinically ascertained sample was 46 (SD=15) and 51% were male. In the UK Biobank a total of 1453 participants had a schizophrenia or SA-D diagnosis, of which 1180 had a medical record diagnosis, 708 had a self-reported diagnosis, and 450 had both. In the UK Biobank sample, the mean age of recruitment was 57 (SD=8) and 46% were male.

### Positive predictive values

In the clinically ascertained sample, a total of 365 participants had both a self-reported diagnosis and a research interview diagnosis. Table 2 details the PPV values. For participants who self-reported a lifetime clinical diagnosis of schizophrenia the PPV of receiving a research interview diagnosis of schizophrenia was 0.70. This increased to 0.81 when benchmarked against a research interview diagnosis of either a schizophrenia or SA-D, and 0.87 when benchmarked against a research interview diagnosis of either schizophrenia, SA-D, or other psychotic disorder. The PPV of self-reporting psychosis (without also reporting schizophrenia or bipolar) and receiving a schizophrenia, SA-D, or other psychotic disorder research interview diagnosis ranged from 0.27-0.65 (Supplementary Table 3).

**Table 2.** Positive predictive values of self-reported diagnoses and subsequent research interview diagnoses.

| Self-report method | Self-reported diagnosis | Research interview diagnosis | Number of participants who self-reported schizophrenia | Number of participants who subsequently met research interview diagnosis | PPV |
| --- | --- | --- | --- | --- | --- |
| **Lifetime clinical diagnosis** | Schizophrenia | Schizophrenia | 273 | 190 | 0.70 |
|  | Schizophrenia | Schizophrenia/SA-D | 273 | 222 | 0.81 |
|  | Schizophrenia | Schizophrenia/SA-D/other psychotic disorders | 273 | 238 | 0.87 |
| **Current clinical diagnosis** | Schizophrenia | Schizophrenia | 239 | 176 | 0.74 |
|  | Schizophrenia | Schizophrenia/SA-D | 239 | 202 | 0.85 |
|  | Schizophrenia | Schizophrenia/SA-D/other psychotic disorders | 239 | 215 | 0.90 |
| **Participant opinion** | Schizophrenia | Schizophrenia | 102 | 77 | 0.75 |
|  | Schizophrenia | Schizophrenia/SA-D | 102 | 84 | 0.82 |
|  | Schizophrenia | Schizophrenia/SA-D/other psychotic disorders | 102 | 87 | 0.85 |

*PPV, positive predictive value; SA-D, schizoaffective disorder depressive-type.*

A self-report of having a current clinical diagnosis of schizophrenia had higher PPVs than both self-reported lifetime clinical diagnosis and self-reported participant’s opinion, across all diagnostic categories. All participants who self-reported schizophrenia but did not proceed to get a research interview diagnosis of schizophrenia received other mood or psychotic research diagnoses, except one participant where there was insufficient data to form a research interview diagnosis (Supplementary Table 4).

In UK Biobank, the PPV of having a medical record diagnosis of schizophrenia in those who self-reported a schizophrenia diagnosis was 0.74 (n=450), which increased to 0.80 when including a medical record diagnosis of schizophrenia or any other psychotic related disorder (n=491). The specificity values were high (0.99953 & 0.99962), demonstrating a small chance of false positives (Supplementary Table 5 and 6). PPV is influenced by prevalence and the prevalence of schizophrenia in UK Biobank was 0.29%, which is less than the population point prevalence of 0.6%. When correcting for a population prevalence^43^ the PPV increased to 0.83 (See Supplementary Table 5 and 6).

### Phenotypic and genetic differences across diagnosis source

#### Clinically ascertained sample

Next, we compared participants whose *only* source of diagnosis was a self-report of schizophrenia (n=654) to participants who had a research interview diagnosis of schizophrenia or SA-D (n=458) in NCMH participants. Participants who had a research interview diagnosis were younger (mean age 43 vs 47; OR = 0.77; 95% CI= 0.67, 0.88; p=9.11×10^−5^) and more likely to have a high school qualification (GCSE) or above (OR = 1.61; 95% CI= 1.13, 2.29; p=0.008) than self-report only participants. Having a degree did not significantly differ across self-report and research interview groups (OR=1.15, 95%CI= 0.79, 1.67, p=0.47). No significant differences were detected in employment (OR = 1.35; 95% CI= 0.87, 2.08; p=0.18) or sex (OR = 1.07; 95% CI= 0.83, 1.37; p=0.61) (Figure 1).

**Figure 1.**
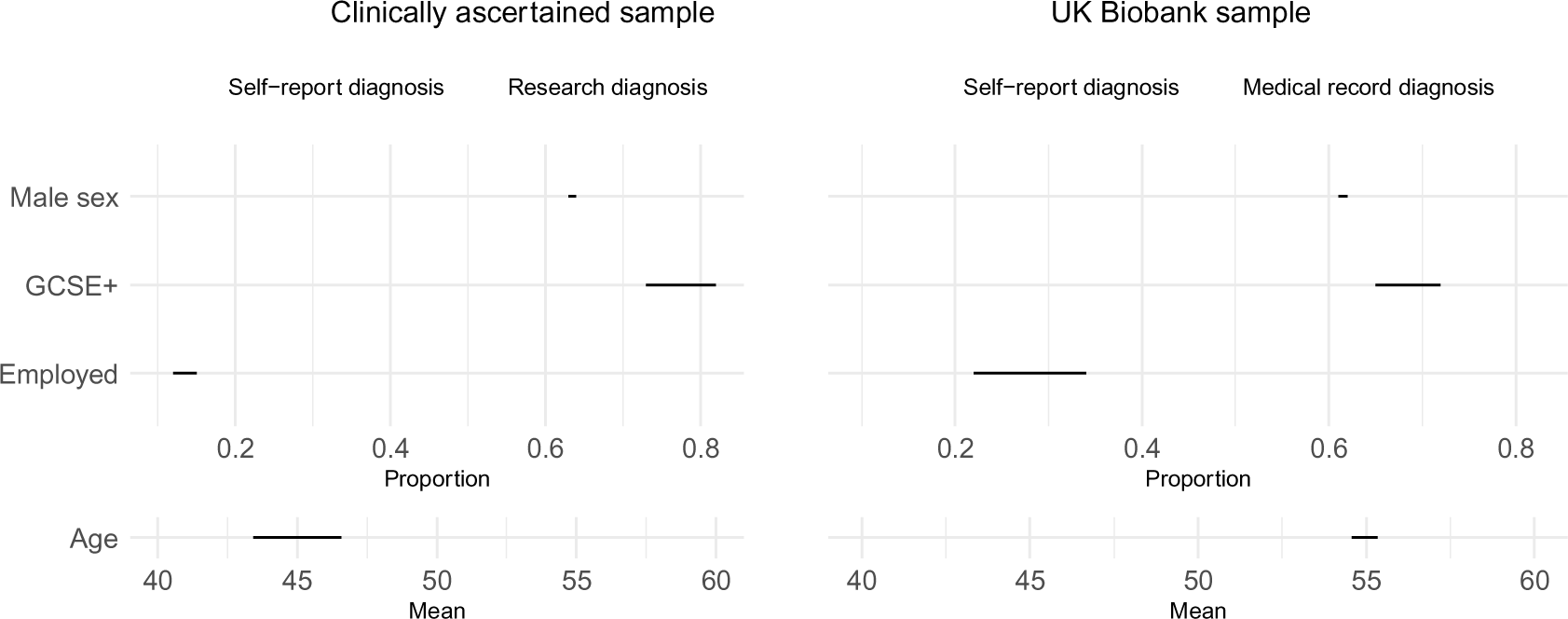
Phenotype differences across methods of diagnosis. Cleveland plot of the proportion of participants that were male, had a GCSE qualification or above, were employed, and mean age by those who had a self-report diagnosis only and a research interview diagnosis/medical record diagnosis.

Across NCMH and CardiffCOGS participants, we found no significant difference in schizophrenia PRS between participants who had a research interview diagnosis (n=803) and those who only self-reported a diagnosis (n=449) (OR = 0.97; 95% CI= 0.86, 1.09; p=0.59) (Figure 2).

**Figure 2.**
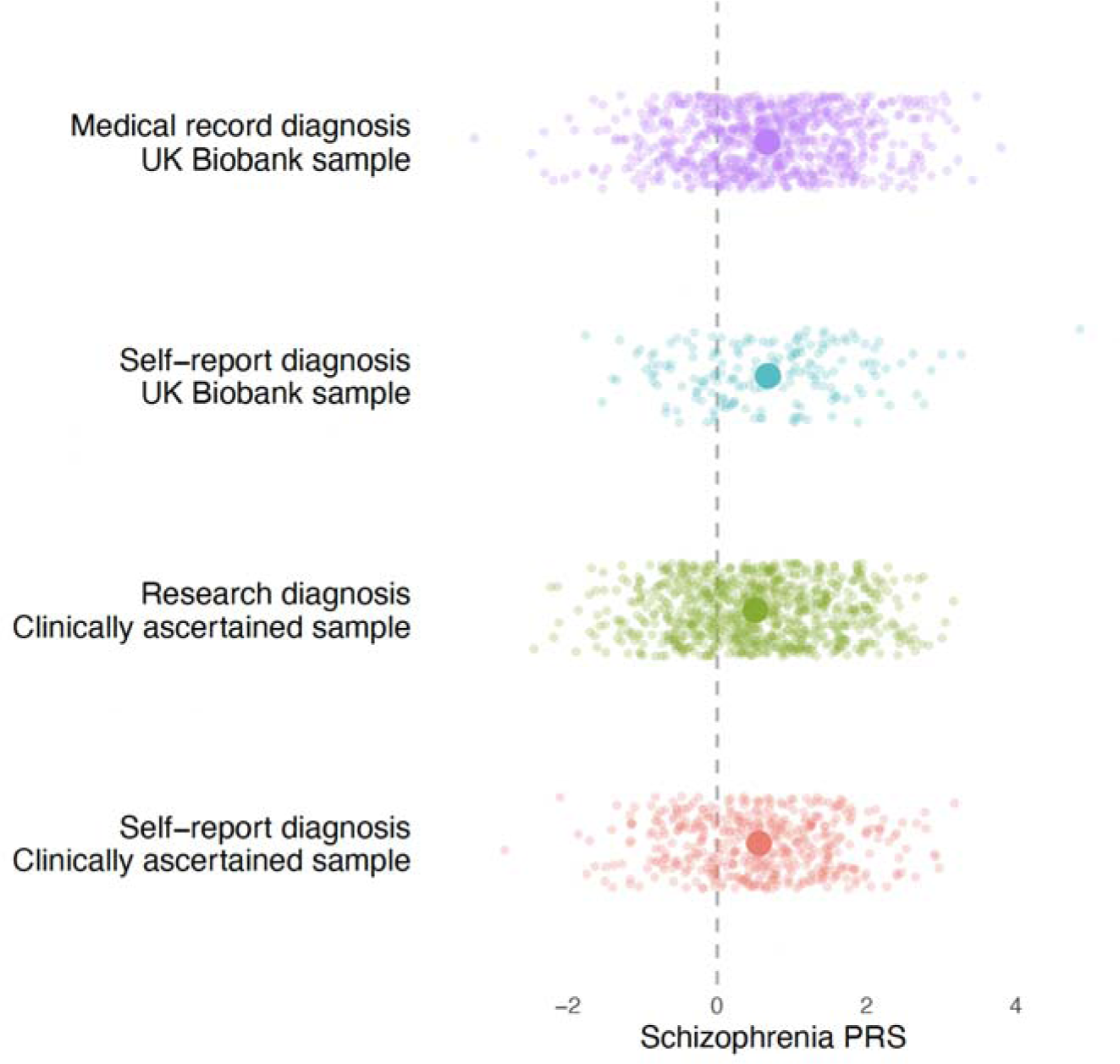
Schizophrenia polygenic risk scores plotted by method of diagnosis. Standardised polygenic risk scores for each method of diagnosis across both samples. The bold circles represent the mean of each diagnostic group. PRS; polygenic risk score.

#### UK Biobank sample

Compared to participants whose basis for a diagnosis was solely self-report (n=252), participants who had a medical record diagnosis of schizophrenia or SA-D (n=1201) were less likely to be in paid employment (OR = 0.55; 95% CI= 0.39, 0.79; p=0.001), and less likely to have a GCSE or above (OR = 0.70; 95% CI=0.51, 0.95; p=0.02). Furthermore, participants with a medical record diagnosis were less likely to have a degree (OR=0.59, 95%CI = 0.44, 0.79, p=0.0005). There were no differences in sex (OR = 0.95; 95% CI=0.72, 1.26; p=0.75) or age across the groups (OR = 0.91; 95% CI= 0.80, 1.04; p=0.18) (Figure 1).

No significant difference in schizophrenia PRS was found between participants who had a medical record diagnosis (n=809) and a self-report diagnosis (n=181) (OR = 1.01; 95%CI= 0.87,1.19; p=0.85) (Figure 2).

### Liability explained in case/control status

The proportion of variance on the liability scale attributable to schizophrenia PRS in both diagnostic groups in the clinically ascertained sample and UK Biobank studies fell within the distribution of studies in the latest PGC analysis (Figure 3, Supplementary Table 8). In the clinically ascertained sample, the schizophrenia PRS explained 5.0% of the variability in the self-report-only group, and 4.7% in the research interview diagnosis group. In the UK Biobank sample, the schizophrenia PRS explained 6.5% of the variability in the self-report only-group and 6.1% in the medical record diagnosis group.

**Figure 3.**
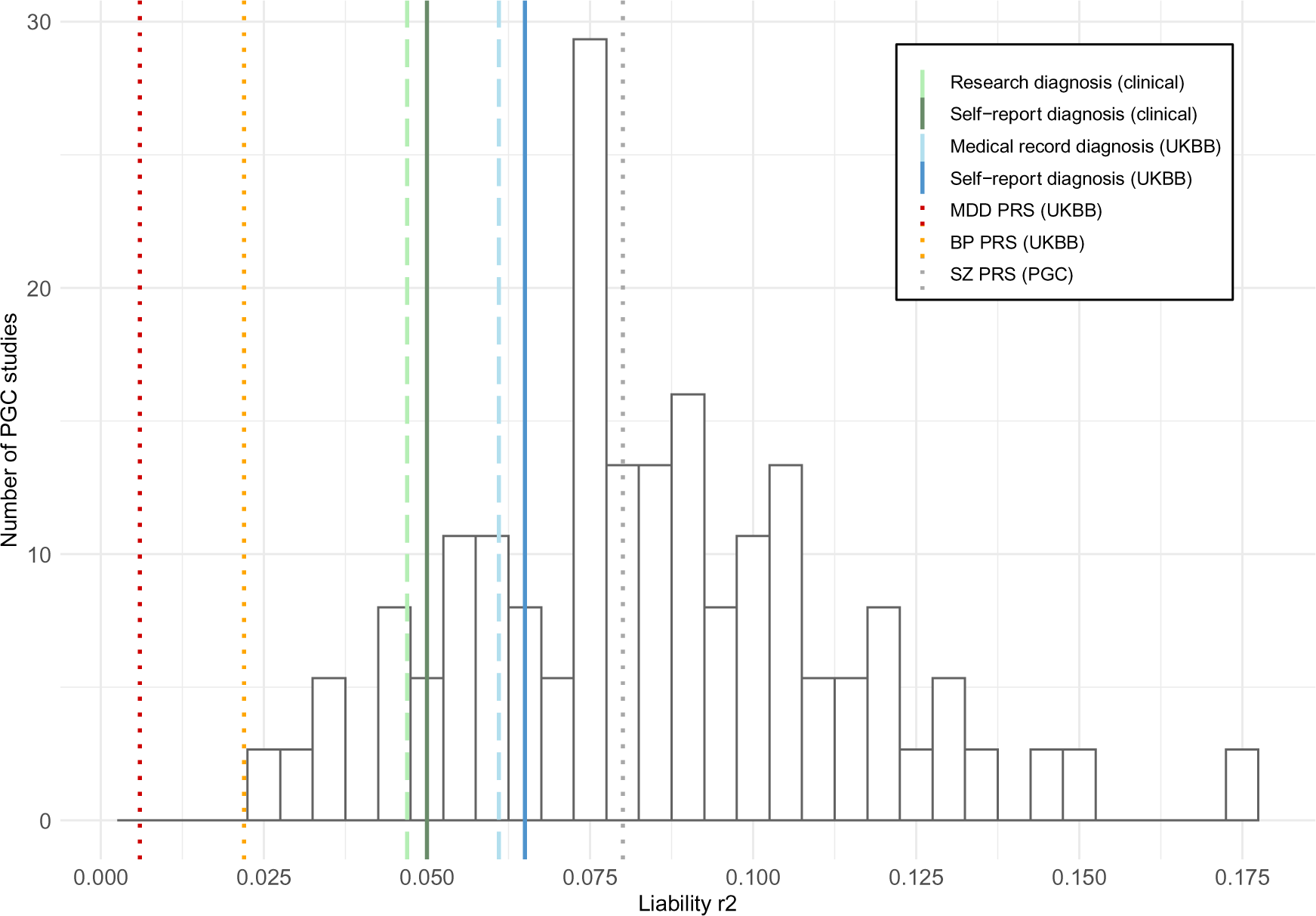
Variance explained by schizophrenia PRS by diagnostic method compared to PGC studies. The variance explained by schizophrenia PRS in schizophrenia case/control status on the liability scale assuming a 1% population prevalence of European genetic ancestry PGC cohorts. The lines plotted on the graph represent the r^2^ of each diagnostic group. SZ, MDD and BP PRS are plotted as a reference. Clinical; clinically ascertained sample, UKBB; UK Biobank, MDD; major depressive disorder, BP; bipolar disorder, SZ; schizophrenia, PGC; Psychiatric Genomics Consortium.

### Further examination of diagnosis source in the UK Biobank

Schizophrenia PRS increased with the number of times a schizophrenia diagnosis was reported; OR=1.82 (95%CI = 1.67,1.99) for 1 endorsement compared to controls and OR= 2.11 (95%CI=1.92,2.32) for 2 or more endorsements (Supplementary Figure 5). Participants who had two or more diagnosis endorsements had a significantly higher schizophrenia PRS than participants who only had one diagnosis endorsement (OR=1.15, 95%CI=1.01,1.31, P=0.03). The schizophrenia PRS also increased as the number of schizophrenia hospital admissions increased; OR=1.85 (95%CI = 1.72,2.00) for 0 admissions (alternative source of schizophrenia diagnosis), OR=1.92 (95%CI = 1.77,2.08) for 1 admission, OR=2.28 (95%CI = 2.01,2.58) and for 2 or more admissions (Supplementary Figure 6).

Schizophrenia cases with a primary ICD-10 admission code had a higher schizophrenia PRS than those who had schizophrenia as a secondary ICD-10 admission code (OR=1.28, 95%CI=1.10,1.49, P=0.002). Participants identified with a schizophrenia diagnosis from a secondary code only, on average, had lower schizophrenia PRS than those identified from self-report or a primary hospital admission code (Supplementary Figure 7). These findings did not appear to be related to the secondary code only participants having different associated diagnoses (Supplementary Table 9 and 10).

## Discussion

The results demonstrated that participants who self-reported a clinical diagnosis of schizophrenia were likely to be given a subsequent research interview diagnosis of schizophrenia, SA-D or other psychotic disorder (PPVs between 0.70 and 0.90). Furthermore, participants in UK Biobank who self-reported a clinical schizophrenia diagnosis were likely to have a medical record diagnosis of schizophrenia or another psychotic disorder (PPV=0.80). Although we found some phenotypic differences, genetic liability to schizophrenia did not significantly differ between participants with a self-reported diagnosis compared to those diagnosed via research interview or medical records. The variance explained by the schizophrenia PRS for all diagnostic methods fell within the distribution of PGC studies. These findings suggest that using a self-reported clinical diagnosis of schizophrenia is a valid approach for identifying participants for large-scale genomic research.

In the clinically ascertained sample, participants who self-reported schizophrenia were likely to receive a research diagnosis of schizophrenia, SA-D or other psychotic disorder, however, participants who self-reported a lifetime clinical diagnosis of psychosis (without schizophrenia and bipolar) were much less likely to obtain a research interview diagnosis of schizophrenia, SA-D, or other psychotic disorder (PPVs 0.27-0.65). Previous research has shown that a schizophrenia diagnosis has much better agreement between diagnostic methods (PPVs 0.69-1.00) than other diagnoses such as bipolar, depression and other psychotic disorders^9^. Although self-reported diagnoses have generally been shown to have poor predictive accuracy when it comes to obtaining a gold-standard research interview diagnosis ^44, 45^, our results suggest that for schizophrenia specifically, self-reported diagnoses could be used in place of a research interview diagnosis to identify participants in research.

In UK Biobank, participants who self-reported schizophrenia were likely to have a medical record diagnosis of schizophrenia. However, the low sensitivity values indicate that a self-report in the UK Biobank did not capture everyone who had a medical record. This could be for many different reasons including later onset of illness, the stigma associated with reporting a schizophrenia diagnosis, or due to the way the question was asked in the assessment; prompts, if any, were only given for physical health conditions and mental health was not specifically mentioned. The high negative classifications in UK Biobank illustrated that participants who did not self-report schizophrenia also did not have a medical record of schizophrenia and vice versa, demonstrating that self-reported diagnoses are effective at ruling out non-cases.

Currently, using a research interview and note review to obtain a diagnosis is considered gold standard, although we find the requirement for detailed and time-consuming interviews may induce recruitment biases, with those participating in such an interview (after the majority having a brief interview first) being younger and more likely to have a high school qualification (GCSEs) than those who only have self-reported. No difference in degree qualification was observed across groups, however this is likely due to the small proportion of individuals with a degree in the clinical sample (research interview=18% vs self-report=16%). Research interviews may exclude participants who are more acutely unwell or cognitively impaired and unable to complete a long assessment. In the UK Biobank, participants with a medical record diagnosis were less likely to have GCSEs and to be employed. Furthermore, participants with a medical record diagnosis were increasingly less likely to have a degree (medical record = 25% vs self-report = 36%). This suggests that these participants may have more impaired functioning than those with a self-report only, and by using medical records only as researchers we may be missing participants who are functioning well and/or have not been admitted to hospital. By using alternative methods, such as a self-report of a diagnosis made by a health professional, research participants may be more representative of people with schizophrenia, although this comes at a cost of diagnostic accuracy.

In contrast to the depression study in UK Biobank which found participants defined by minimal phenotyping (self-report, help seeking, and symptom based) had lower SNP-derived heritability than the strictly defined participants (Composite International Diagnostic Interview) ^23^, we did not find a difference in schizophrenia PRS between the self-report and research interview/medical record diagnosis groups. This highlights the potential, especially for genomic studies, of using this method to identify participants for schizophrenia research. However, we did find differences in schizophrenia PRS within hospital admission diagnoses (in primary and secondary admissions). We also found the number of diagnosis reports and admissions were associated with higher polygenic risk scores in UK Biobank, as has been reported in previous literature^46, 47^. This is consistent with findings from the PGC, who reported schizophrenia PRS to be higher in patients who were recruited from inpatient settings^27^. These findings could indicate greater severity or improved accuracy of diagnosis, or both. We also found participants whose primary reason for admission was schizophrenia, and those that only self-reported schizophrenia, had a higher schizophrenia PRS than those with a secondary admission diagnosis. One explanation of the difference in schizophrenia PRS could be that the secondary admission group were participants who were not admitted primarily for psychosis because their symptoms were milder or were well treated. Alternatively, the accuracy of a secondary diagnosis may be more prone to error than a diagnosis given for a primary admission to hospital (e.g., if admitted for a heart attack), although this did not appear to be the case when looking specifically at psychiatric comorbidities.

### Limitations

It is important to note limitations of the current study. Participants were invited to complete a SCAN-based research interview if they self-reported psychosis or a schizophrenia diagnosis. This study design prevented us from assessing other metrics (negative predictive value, sensitivity, and specificity) in the clinically ascertained sample. This also meant we were unable to adjust the PPV to the population point prevalence of schizophrenia. As a result, the PPV could have been inflated by the high proportion of schizophrenia participants in our clinically ascertained sample. Additionally, some participants had their diagnosis confirmed by a clinician if systematically recruited, which could have increased the positive predictive values, and only a subset of the sample were asked their own opinion of their diagnosis (n=99). Despite these limitations, our clinically ascertained sample is one of the only psychosis-based samples with both self-report diagnosis data and a gold-standard research diagnosis. In the UK Biobank, 93% of the participants with a medical record diagnosis of schizophrenia have a hospital admission, therefore the predictive values primarily reflect how predictive a self-report was of a hospital admission.

Both the clinically ascertained sample and UK Biobank were sampled from the UK; therefore, the findings may not apply to other countries. The generalisability of the UK Biobank findings are also hindered because this sample is not wholly representative of the UK population^48^ and the data could have favoured younger participants as diagnoses were drawn from electronic medical records rather than paper records which may not have been transferred. Furthermore, the primary and secondary admission diagnosis in the UK Biobank may have differed depending on the location of the admission (e.g., psychiatric hospital vs general) or by the clinician’s expertise. Polygenic risk scores were restricted to participants from a European genetic ancestry. We were unable to investigate whether the polygenic risk scores differ across diagnostic groups in non-European genetic ancestries due to a limited number of participants in our samples from non-European genetic ancestries. Lastly, the recruitment methods in the NCMH study could have enriched for relatives of those with mood and psychotic disorders, although only 5% (n=33) of our controls reported having a family history of bipolar disorder or schizophrenia. Nonetheless, and although the effect of this would be conservative, this could weaken the variance explained in our schizophrenia PRS analyses.

## Conclusion

Self-reporting a clinical schizophrenia diagnosis may be a valid method for identifying cases in schizophrenia research, providing systematic differences of methodologies are transparently noted. Participants who only self-reported a schizophrenia diagnosis showed differences in age, education, and employment but they do not differ in relation to schizophrenia genetic liability. These findings provide preliminary evidence for using less stringent methods of ascertaining diagnoses in schizophrenia research, particularly in genetic research, which could in turn improve the representativeness of future samples and increase sample sizes.

## Data Availability

UK Biobank data is available upon application to them. NCMH data is available upon request.

## Acknowledgements

We thank the participants, clinicians, lab staff and field team for their help with the NCMH, CardiffCOGS and UK Biobank studies.

## Conflict of Interest

JTRW, MCO’D and MJO received a research grant to Cardiff University from Takeda Pharmaceuticals that funded this work and GW’s research position. Takeda Pharmaceuticals have not had any input into the study design, analysis, or interpretation of results. JTRW, MCO’D, MJO, IRJ and AFP have received research funding from Akrivia Health for work unrelated to this study.

## Funding

JTRW, MCO’D and MJO received a research grant to Cardiff University from Takeda Pharmaceuticals that funded this work and GW’s research position. This work was supported by the following grants: Medical Research Council Program (MR/P005748/1), DATAMIND (MR/W014386/1) and a grant from NIH (Award U01MH109514).

## Notes

### Competing Interest Statement

The authors have declared no competing interest.

### Funding Statement

JTRW, MCOD and MJO received a research grant to Cardiff University from Takeda Pharmaceuticals that funded this work and GWs research position. This work was supported by the following grants: Medical Research Council Program (MR/P005748/1), DATAMIND (MR/W014386/1) and a grant from NIH (Award U01MH109514).

### Author Declarations

NCMH and CardiffCOGS received approval from Health Research Authority and Wales Research Ethics Committee (REC) 2 (NCMH REC reference: 16/WA/0323), and Southeast Wales REC (CardiffCOGS REC reference: 07/WSE03/110). All participants provided written informed consent. Ethical approval was granted to the UK Biobank by the Northwest Multi-Centre Ethics Committee. This study was conducted under UK Biobank project number 13310.

