## Supplementary Materials for "Assessing the validity of a self-reported clinical diagnosis of schizophrenia"

### Table 1. Field IDs for UK Biobank diagnosis definitions

| **UK Biobank psychotic disorders diagnosis** | **Field ID** |
| --- | --- |
| *Medical record diagnosis* | |
| Primary care diagnosis | 130875 (code 30/31) |
| Hospital admission | 130875/130885 (code 40/41) |
| - Primary hospital admission | 41202 |
| - Secondary hospital admission | 41204 |
| Death records | 130875 (code 20/21) |
| *Self-report diagnosis* | |
| Mental health questionnaire | 20544 (code 2) |
| Verbal self-report | 20002 (code 1289) |

Table 1 outlines UK Biobank field IDs used to create diagnosis groups.

### Table 2. Number of participants per diagnostic method in the phenotypic and genetic differences analysis.

| **Sample** | **Population** | **Diagnostic method in cases** | **Number of participants per diagnostic method (NCMH)** | **Number in genetic analyses (NCMH & CardiffCOGS)** |
| --- | --- | --- | --- | --- |
| **Clinical sample** | Clinical population | Self-report | 654 | 449* |
|  |  | Research interview diagnosis | 458 | 803* |
| **UK Biobank** | General population-based | Self-report | 252 | 181* |
|  |  | Medical record diagnosis | 1201 | 809* |

Table 2 shows the number of participants per diagnostic method and number of participants in the genetic analysis by self-report only/research interview diagnosis (clinically-ascertained sample) and self-report only/medical record diagnosis (UK Biobank). *Participants of European genetic ancestry.

### Table 3. Positive predictive values of psychosis in the clinically-ascertained sample

| Self-report method | Self-reported diagnosis | Research diagnosis | Number of participants who self-report psychosis (without schizophrenia/bipolar) | Number of participants who subsequently met research interview diagnosis | PPV |
| --- | --- | --- | --- | --- | --- |
| Lifetime clinical diagnosis | Psychosis | Schizophrenia | 117 | 33 | 0.28 |
|  | Psychosis | Schizophrenia/SA-D | 117 | 58 | 0.50 |
|  | Psychosis | Schizophrenia/SA-D/other psychotic disorders | 117 | 71 | 0.61 |
| Current clinical diagnosis | Psychosis | Schizophrenia | 94 | 25 | 0.27 |
|  | Psychosis | Schizophrenia/SA-D | 94 | 45 | 0.48 |
|  | Psychosis | Schizophrenia/SA-D/other psychotic disorders | 94 | 58 | 0.62 |
| Participant opinion | Psychosis | Schizophrenia | 51 | 22 | 0.43 |
|  | Psychosis | Schizophrenia/SA-D | 51 | 27 | 0.53 |
|  | Psychosis | Schizophrenia/SA-D/other psychotic disorders | 51 | 33 | 0.65 |

Table 3 shows the positive predictive values (PPV) of individuals who self-reported psychosis (without also reporting schizophrenia and bipolar disorder) and subsequent research interview diagnoses.

### Table 4. Diagnoses of participants who did not receive a schizophrenia or SA-D research diagnosis

| **DSM 4 Diagnosis** | **Number of participants** |
| --- | --- |
| Alcohol induced psychosis | 2 |
| Bipolar disorder type 1 | 8 |
| Brief psychotic disorder | 2 |
| Cyclothymia | 2 |
| Delusional disorder | 1 |
| Major depressive disorder recurrent | 5 |
| Major depressive disorder single episode | 1 |
| Psychosis not otherwise specified | 8 |
| Psychotic depression | 4 |
| Schizoaffective bipolar type | 15 |
| Substance induced psychotic disorder | 2 |
| Unknown | 1 |

Table 4 shows the DSM 4 diagnoses of participants who self-reported schizophrenia and did not receive a SCAN-based research interview diagnosis of schizophrenia or schizoaffective disorder depressive type (SA-D) (n=51).

### Table 5. Predictive values of participants who self-report a schizophrenia diagnosis and have a medical record diagnosis of schizophrenia in the UK Biobank.

|  | **Medical record schizophrenia**  **Yes** | **Medical record schizophrenia**  **No** |  |
| --- | --- | --- | --- |
| **Self-reported schizophrenia**  **Yes** | 450 | 156 | PPV: 0.7425743 |
| **Self-reported schizophrenia**  **No** | 724 | 333784 | NPV: 0.9978356 |
|  | Sensitivity: 0.3833049 | Specificity: 0.9995329 |  |

Table 5 shows the positive predictive values (PPV), negative predictive values (NPV), sensitivity and specificity of individuals who self-reported schizophrenia either verbally to a nurse on the initial assessment or on the mental health questionnaire and had a medical record diagnosis of schizophrenia. Adjusted values based on point prevalence:

PPV: (0.3833049*0.006)/((0.3833049*0.006)+((1-0.9995329)*(1-0.006))) = 0.8320275

NPV: (0.9995329*(1-0.006))/((0.9995329*(1-0.006))+((1-0.3833049)*0.006)) = 0.9962896

### Table 6. Predictive values of participants who self-report a schizophrenia diagnosis and have a medical record diagnosis of schizophrenia or other psychotic disorders in the UK Biobank.

|  | **Medical record schizophrenia**  **/other psychotic disorders**  **Yes** | **Medical record schizophrenia**  **/other psychotic disorders**  **No** |  |
| --- | --- | --- | --- |
| **Self-reported schizophrenia**  **Yes** | 491 | 124 | PPV: 0.798374 |
| **Self-reported schizophrenia**  **No** | 1853 | 332852 | NPV: 0.9944638 |
|  | Sensitivity: 0.209471 | Specificity: 0.9996276 |  |

Table 6 shows the positive predictive values (PPV), negative predictive values (NPV), sensitivity and specificity of individuals who self-reported schizophrenia either verbally to a nurse on the initial assessment or on the mental health questionnaire and had a medical record diagnosis of schizophrenia or other psychotic disorders. Other psychotic disorders include codes corresponding to schizotypal disorder, persistent delusional disorders, acute and transient psychotic disorders, induced delusional disorder, schizoaffective disorders, other nonorganic psychosis and unspecified nonorganic psychosis. Adjusted values based on point prevalence:

PPV: (0.209471*0.006)/((0.209471*0.006)+((1-0.9996276 )*(1-0.006))) = 0.7724846

NPV: (0.9996276 *(1-0.006))/((0.9996276 *(1-0.006))+((1-0.209471)*0.006)) = 0.9952491

### Table 7. Number of participants per genotyping array

| **Array** | **Number of CardiffCOGS cases** | **Number of NCMH cases** | **Number of NCMH controls** |
| --- | --- | --- | --- |
| GSA | 0 | 981 | 484 |
| OmniExpress | 632 | 0 | 0 |
| PsychArray | 0 | 562 | 265 |

Table 7 shows the number of participants by OmniExpress/PsychArray/GSA array platforms in the genetic subset.

CardiffCOGS samples were genotyped on a different array platform (OmniExpress) to NCMH cases and controls, which were split across GSA and PsychChip (Table 7). To test whether there were any batch effects, we removed the CardiffCOGS samples from the case/control analysis. We found a consistent effect for the PRS predicting schizophrenia case/control status (OR=1.70; 95%CI=1.44-2.02; P=1.91x10^-10^, r^2^=0.036; se=0.011; auc=0.64). This finding, alongside Supplementary Figure 3 suggests that there were no batch effects in the genetic data.

### Table 8. Variance explained by schizophrenia PRS by diagnostic method

|  | Case/control | Definition | Number of participants | OR | 95% CI | P | R^2^ | se | AUC |
| --- | --- | --- | --- | --- | --- | --- | --- | --- | --- |
| ***Clinical sample*** | | | | | | | | | |
| Self-report only | Case | Self-report schizophrenia | 552 | 1.89 | 1.67-2.15 | 7.33x10^-26^ | 0.050 | 0.007 | 0.67 |
|  | Control | Unaffected controls | 710 |  |  |  |  |  |  |
| Research interview | Case | Research interview diagnosis schizophrenia/SA-D | 789 | 1.83 | 1.64-2.05 | 1.02x10^-28^ | 0.047 | 0.007 | 0.66 |
|  | Control | Unaffected controls | 710 |  |  |  |  |  |  |
| ***UK Biobank sample*** | | | | | | | | | |
| Self-report only | Case | Self-report schizophrenia | 494 | 2.01 | 1.84-2.20 | 6.18x10^-53^ | 0.065 | 0.009 | 0.69 |
|  | Control | No schizophrenia or psychotic disorder diagnosis (F20:F29) in first occurrences field (ID=2405) | 401795* |  |  |  |  |  |  |
| Medical record diagnosis | Case | Medical record schizophrenia/SA-D (F20/F25.1) | 809 | 1.96 | 1.83-2.11 | 3.35x10^-80^ | 0.061 | 0.006 | 0.68 |
|  | Control | No schizophrenia or psychotic disorder diagnosis (F20:F29) in first occurrences field (ID=2405) | 401841 |  |  |  |  |  |  |
| ***Other UK Biobank PRS*** | | | | | | | | | |
| Bipolar PRS | Case | Any schizophrenia diagnosis | 990 | 1.50 | 1.41-1.60 | 6.88x10^-37^ | 0.022 | 0.003 | 0.61 |
|  | Control | No schizophrenia, psychotic or mood disorder diagnosis (F20:F39) in first occurrences field (ID=2405) | 352972 |  |  |  |  |  |  |
| Depression PRS | Case | Any schizophrenia diagnosis | 990 | 1.23 | 1.15-1.30 | 1.91x10^-10^ | 0.006 | 0.002 | 0.56 |
|  | Control | No schizophrenia, psychotic or mood disorder diagnosis (F20:F39) in first occurrences field (ID=2405) | 352972 |  |  |  |  |  |  |

*SA-D, schizoaffective disorder depressive-type OR, Odds ratio; 95%CI, 95% confidence intervals; P, p-value; r^2^, variance explained by PRS; se, standard error; AUC, area under the curve.*

Table 8 displays the proportion of variance on the liability scale attributable to schizophrenia PRS in those with a self-reported diagnosis and a SCAN-based research interview diagnosis in the clinically ascertained sample and self-reported diagnosis and medical record diagnosis in UK Biobank. Additionally, Bipolar PRS and Depression PRS in the UK Biobank explaining schizophrenia case/control status are provided as a reference.

### Table 9. Number of other psychotic and mood admissions by primary and secondary schizophrenia admission groups

|  | UK Biobank field ID | Primary schizophrenia admission (0) (N=209) | Secondary schizophrenia admission only (1) (N=459) |
| --- | --- | --- | --- |
| Other psychotic related diagnosis | 130877, 130879, 130881, 130885, 130887, 130889 | 64 (31%) | 115 (25%) |
| Mood diagnosis | 130891:130903 | 73 (35%) | 164 (36%) |

Table 9 shows the number (and percentage) of other psychotic related diagnoses and mood diagnoses of participants who had a primary admission of schizophrenia and a secondary admission of schizophrenia. Diagnoses were assigned based on a hospital admission code (40) or hospital admission and other sources code (41).

(See <https://biobank.ndph.ox.ac.uk/showcase/label.cgi?id=2405> for further details).

### Table 10. Other admission diagnoses by primary and secondary schizophrenia admission groups

|  | UK Biobank field ID | Primary schizophrenia admission (0) (N=93) | Secondary schizophrenia admission only (1) (N=233) |
| --- | --- | --- | --- |
| Delirium | 130847 | 2 (2%) | 25 (11%) |
| Cognitive disorders | 130837:130843 | 3 (3%) | 15 (6%) |
| Substance disorders | 130855:130873 | 12 (13%) | 28 (12%) |
| Anxiety related disorders | 130905:130911 | 16 (17%) | 44 (19%) |
| Other psychiatric diagnosis | 130913:130991 | 15 (16%) | 32 (14%) |

Table 10 shows the number (and percentage) of other mental health diagnoses of participants who did not have either a psychotic related diagnosis or a mood diagnosis by primary and secondary admission groups. Diagnoses were assigned based on a hospital admission code (40) or hospital admission and other sources code (41).

(See <https://biobank.ndph.ox.ac.uk/showcase/label.cgi?id=2405> for further details).

### Figure 1. List of NCMH self-report mental health diagnoses

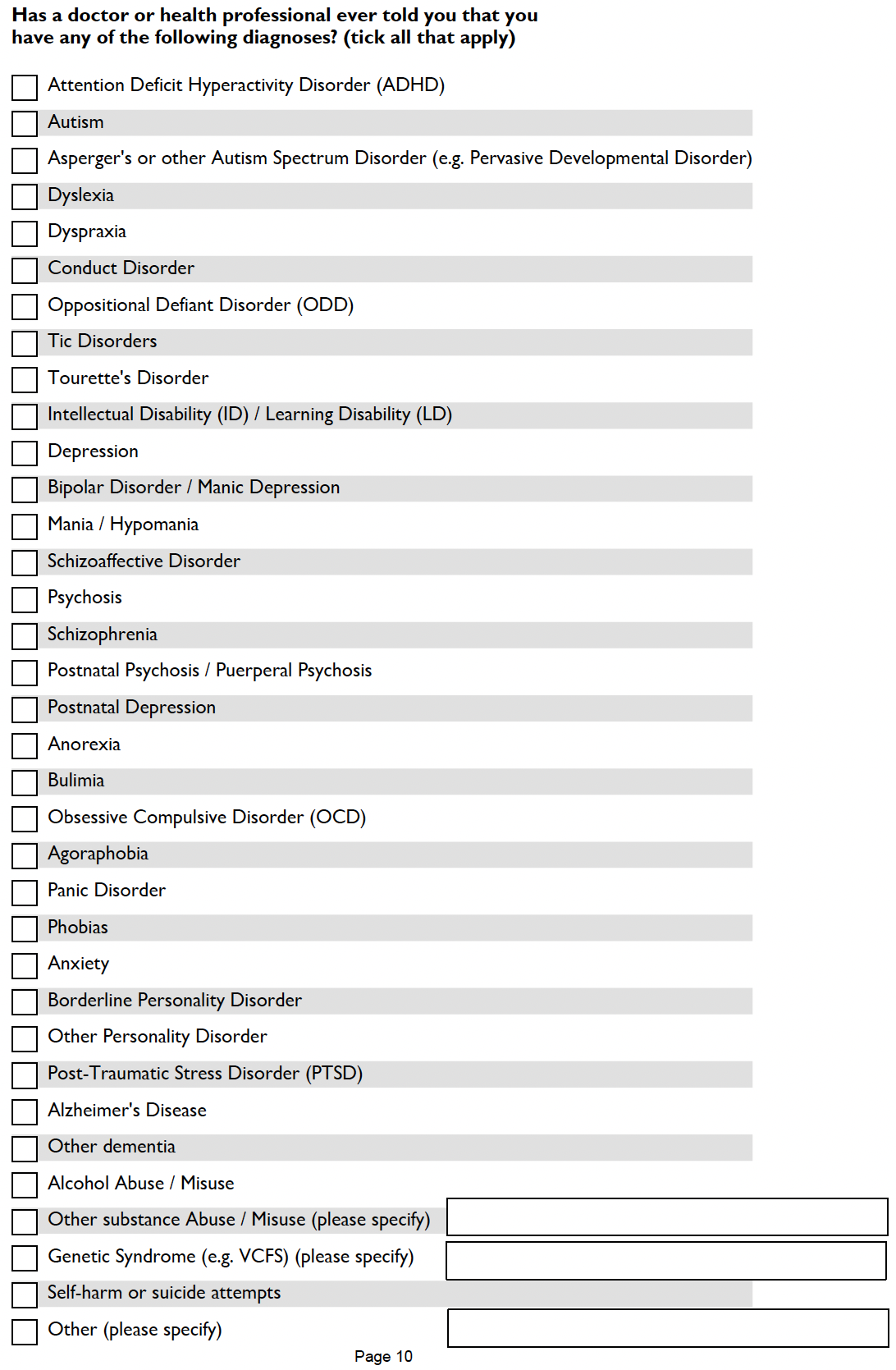

Figure 1 shows a screenshot of the list of mental health diagnoses given to participants to choose from in NCMH.

### Figure 2. Screenshots of self-report mental health diagnoses in the Mental Health Questionnaire in UK Biobank

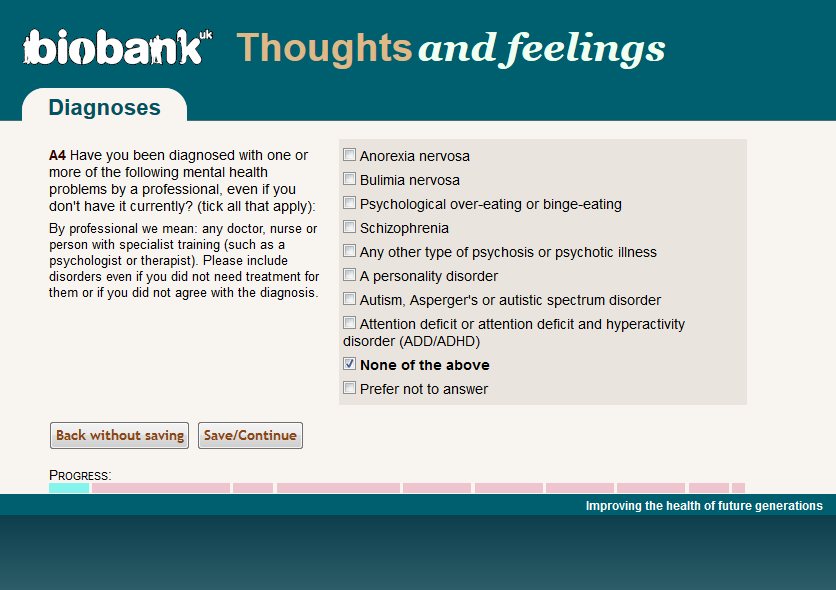

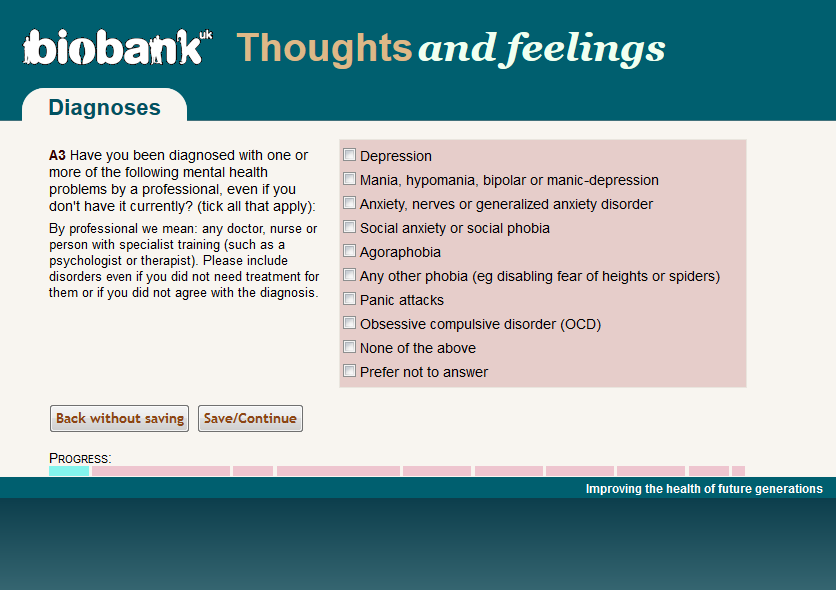
 Figure 2 shows screenshots of the list of mental health diagnoses participants can choose from in the Mental Health Questionnaire in the UK Biobank.

### Figure 3. Plots of principal components 1-6 by array

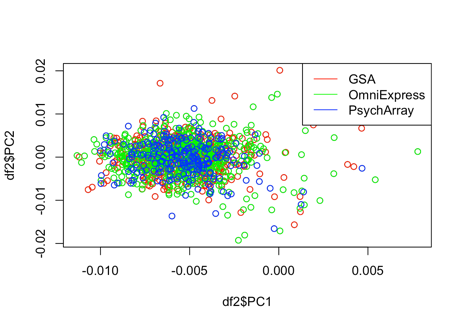

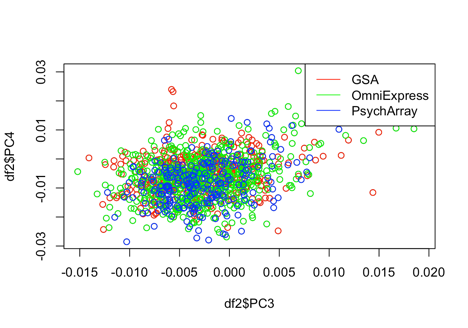

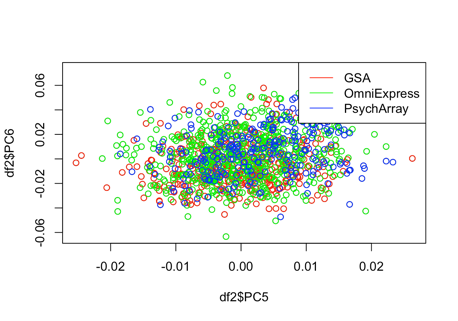

Figure 3 shows principal component plots by each genotyping array (GSA, OmniExpress, PsychArray) to demonstrate any batch effects of the ancestrally homogenous subsample.

### Figure 4. Plots of principal components 1-6 with and without thresholding

a) PC1-6 prior to thresholding

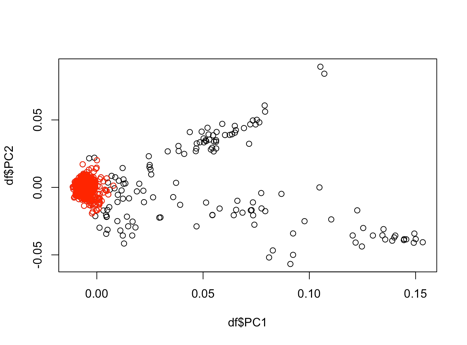

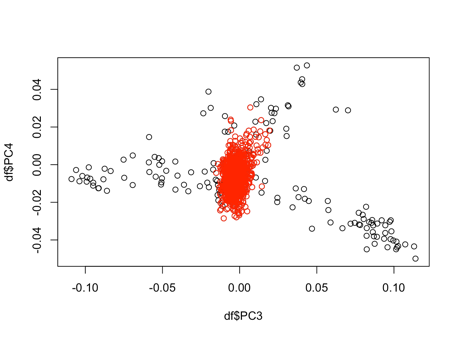

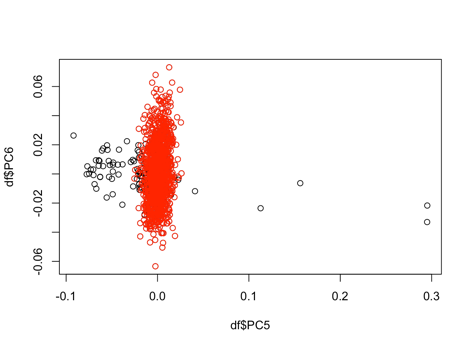

b) PC1-6 of the ancestrally homogenous subset

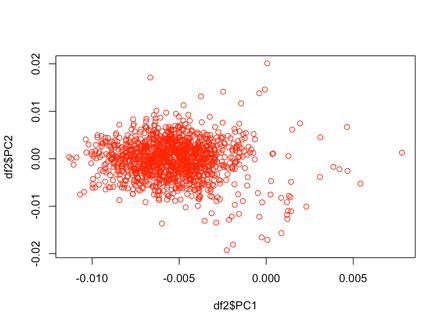

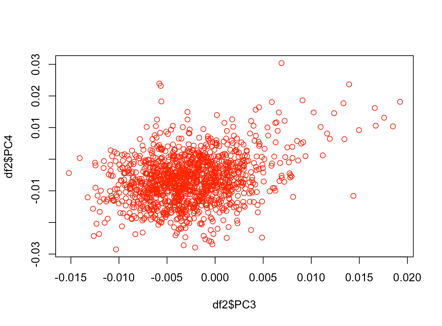

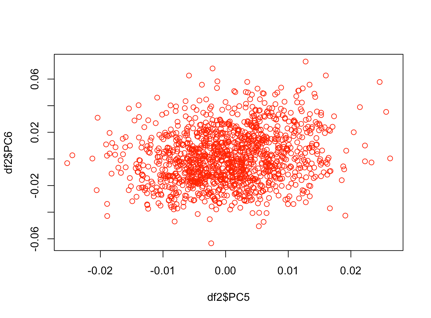

Figure 4 shows the first 6 principal components. The black points represent PRS generated prior to thresholding and the red points highlight the PRS selected to create the ancestrally homogenous subsample. Figure 4b shows an enlarged image of each corresponding PC plot.

### Figure 5. Number of diagnosis endorsement and schizophrenia PRS

**a	b**
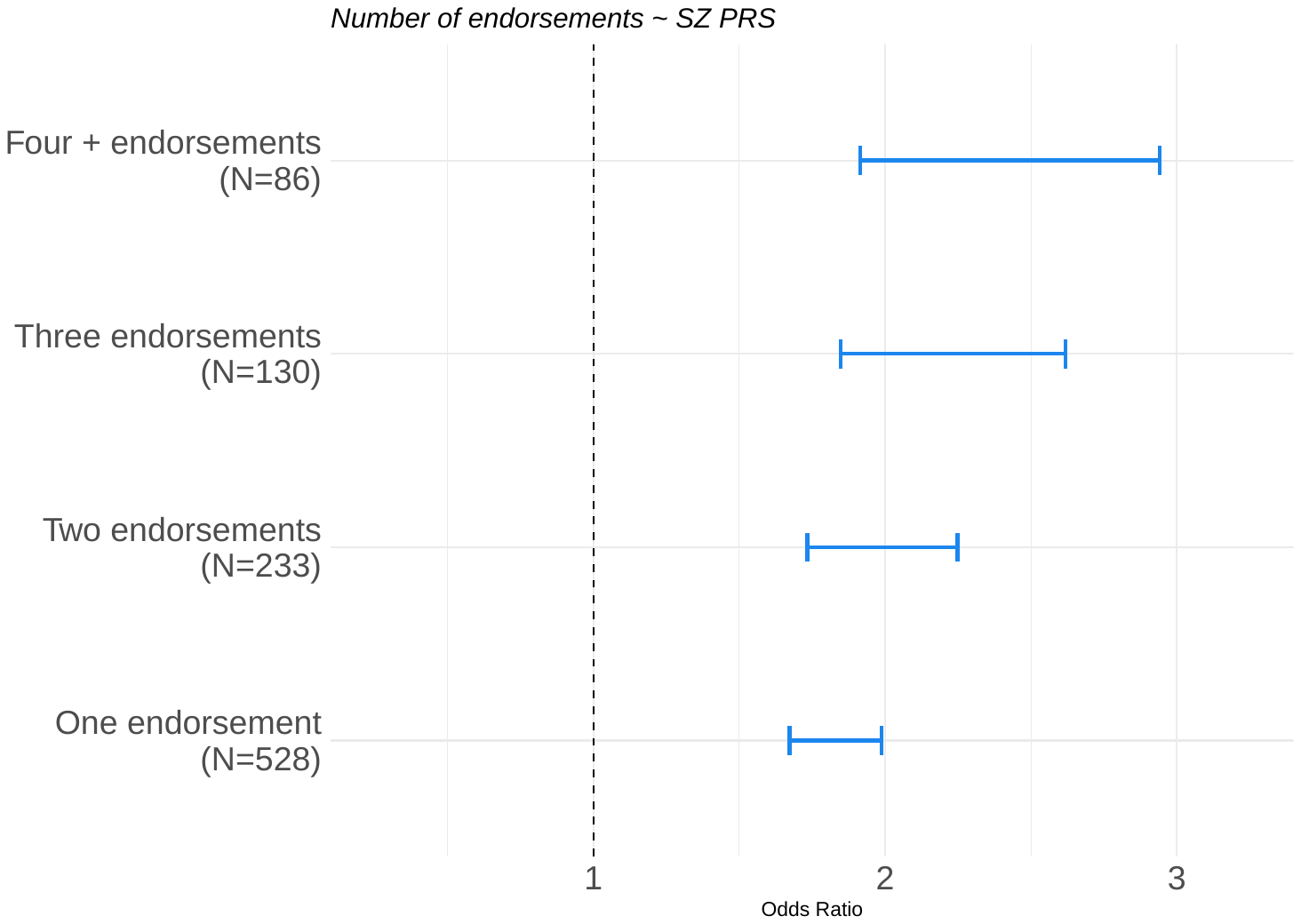

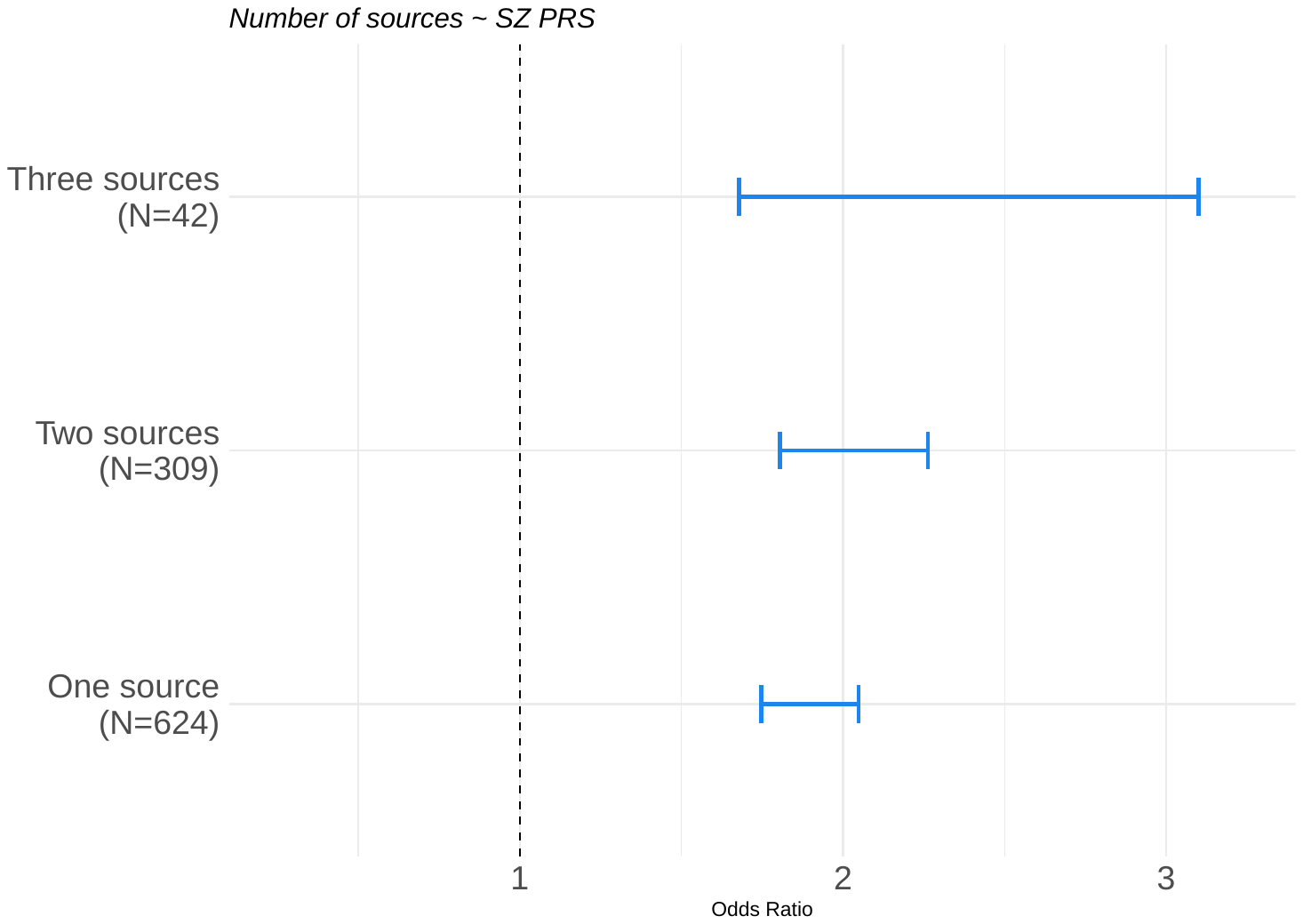

Figure 5 demonstrates as participants’ schizophrenia PRS increased; the number of times a diagnosis was reported increased. Figure 5a displays the number of endorsements (number of times a diagnosis is reported in UK Biobank). Figure 5b shows the same graph when concatenating all hospital admissions into one hospital endorsement.

### Figure 6. Number of schizophrenia admissions and schizophrenia PRS

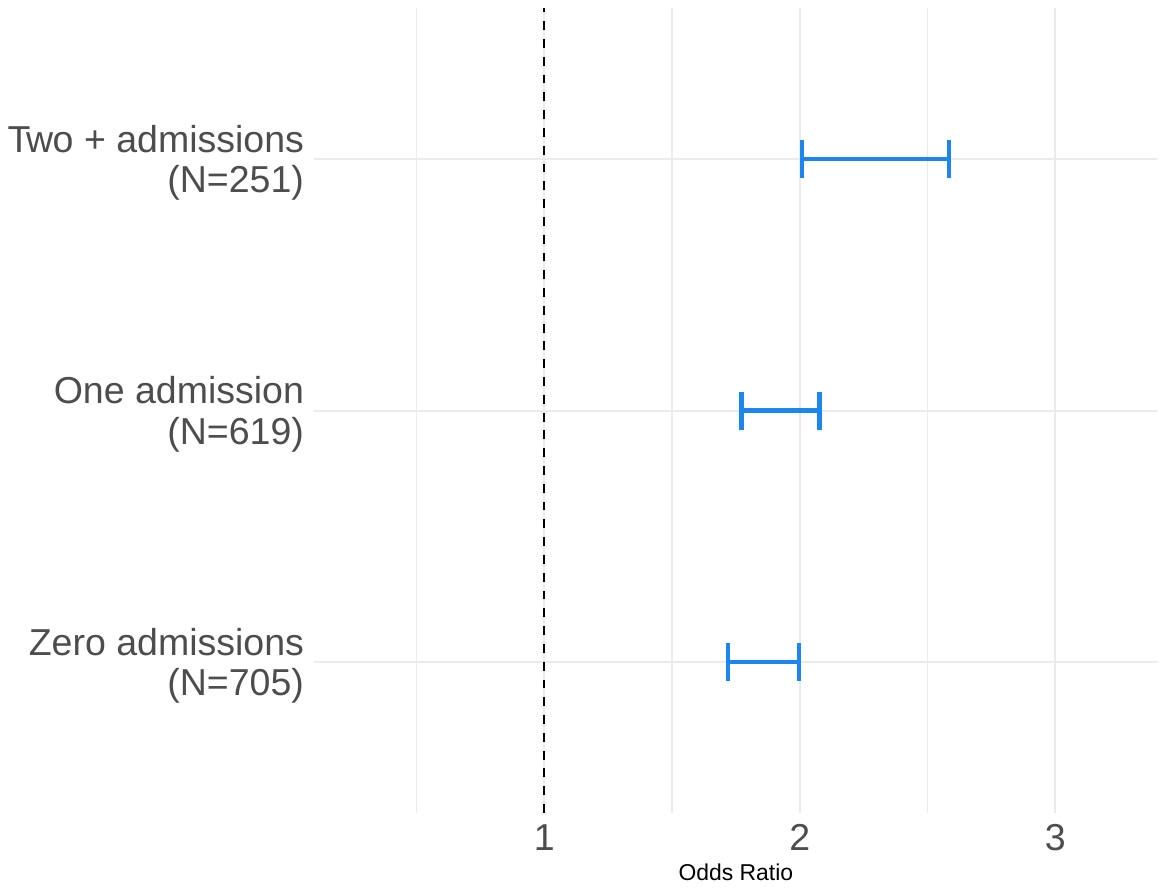

Figure 6 shows as participants schizophrenia PRS increases the odds of having an admission increases.

### Figure 7. Schizophrenia polygenic risk score by primary and secondary admission groups

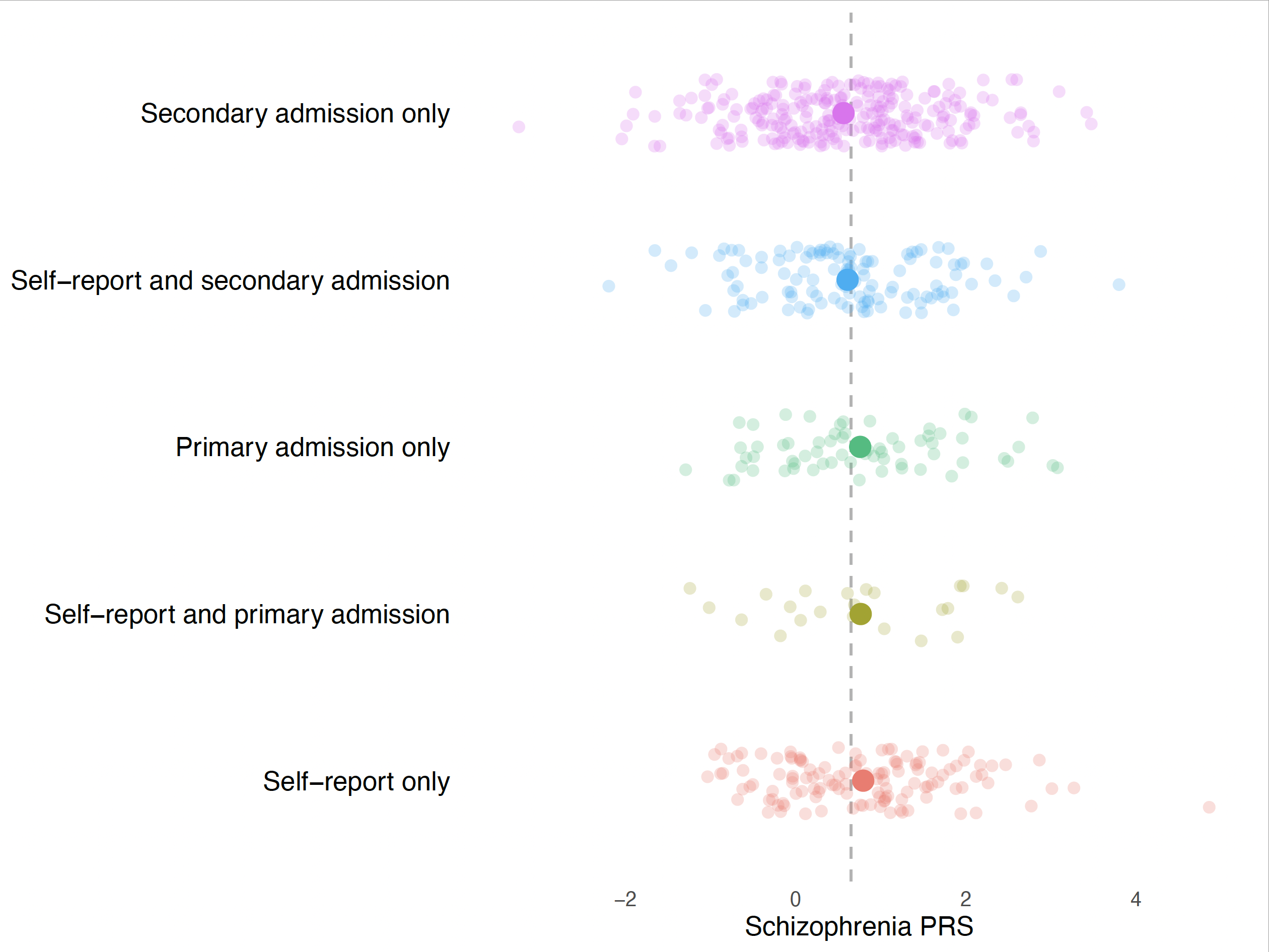

Figure 7 shows the standardised schizophrenia polygenic risk score by primary and secondary admission diagnoses, subdivided into those with only an admission code and those with both a self-report and admission code. The dotted line represents the mean schizophrenia polygenic risk score.

### Supplementary Note. Adjustment for prevalence of schizophrenia.

In the UK Biobank, predictive values were adjusted to the point prevalence of schizophrenia (0.6%). The following formulas were used to adjust the PPV and NPV:

PPV = (sensitivity x prevalence)/[(sensitivity x prevalence)+((1 – specificity) x (1 – prevalence))]

NPV = (specificity x (1 – prevalence))/[(specificity x (1 – prevalence))+((1 – sensitivity) x prevalence)]
